# Cytokine-Driven NF-κB Activation in Retinal Cells and Its Impact on the Pathogenesis of Age-Related Macular Degeneration: A Systematic Review

**DOI:** 10.1101/2024.10.06.24314951

**Authors:** Viqas Shafi, Nabeel Ahmad Khan, Javeria Kazmi, Ifrah Siddiqui

## Abstract

**Objective:** To investigate the role of cytokine-induced NF-κB activation in retinal pigment epithelium (RPE) and photoreceptor cells, elucidating its contribution to the pathogenesis of Age-related Macular Degeneration (AMD) and identifying potential therapeutic targets.

**Background:** AMD is a leading cause of vision loss in older adults, driven by inflammation, oxidative stress, and immune dysregulation, particularly in RPE and photoreceptor cells. The NF-κB signaling pathway, activated by cytokines such as TNF-α, IL-6, and IL-1β, regulates these pathological processes. Understanding cytokine-induced NF-κB activation may provide insight into AMD progression and facilitate the development of novel therapeutic strategies.

**Methods:** A systematic review of literature from databases including PubMed, MEDLINE, and Google Scholar was conducted to evaluate the involvement of TNF-α, IL-6, IL-1β, IL-8, IFN-γ, IL-17, IL-12, IL-18, IL-33, and IL-25 in activating NF-κB in RPE and photoreceptor cells, contributing to AMD pathogenesis. The search was performed with no date restrictions and followed PRISMA guidelines. Eligible studies were selected based on predefined criteria assessing NF-κB activation mechanisms and its effects on inflammation, oxidative stress, and retinal degeneration.

**Results:** The investigation revealed that cytokines TNF-α, IL-6, IL-1β, IL-8, IFN-γ, IL-17, IL-12, IL-18, IL-33, and IL-25 activate the NF-κB signaling pathway in retinal pigment epithelium (RPE) and photoreceptor cells. This activation triggers the transcription of genes involved in inflammation, oxidative stress, and immune responses. Chronic NF-κB activation leads to persistent inflammation, increased oxidative stress, and immune cell recruitment, contributing to retinal cell dysfunction and degeneration. Additionally, NF-κB-driven upregulation of angiogenic factors such as VEGF promotes neovascularization in wet AMD. These findings highlight the central role of NF-κB in AMD pathogenesis and suggest that targeting this pathway could mitigate inflammatory and oxidative damage, offering potential therapeutic benefits for AMD patients.

**Conclusion:** Cytokine-induced NF-κB activation plays a central role in AMD pathogenesis by promoting chronic inflammation, oxidative stress, and angiogenesis in retinal cells. Targeting NF-κB could present a therapeutic strategy to reduce inflammatory and oxidative damage, preserving retinal function and preventing vision loss in AMD. Further research is required to develop effective NF-κB-targeted interventions.

## Background

Age-related macular degeneration (AMD) is a leading cause of vision loss among the elderly, characterized by the progressive degeneration of the retinal pigment epithelium (RPE) and photoreceptor cells. Despite extensive research, the precise molecular mechanisms driving AMD remain incompletely understood. Chronic inflammation, oxidative stress, immune dysregulation, and aberrant angiogenesis are well-recognized contributors to AMD pathology. However, the specific pathways and molecular interactions mediating these processes warrant further investigation [1].

The NF-κB signaling pathway has emerged as a critical regulator of inflammation, oxidative stress, and immune responses. This pathway is activated by various cytokines, including TNF-α, IL-6, IL-1β, IL-8, IFN-γ, IL-17, IL-12, IL-18, IL-33, and IL-25, which are known to play roles in inflammatory and immune responses. Given the centrality of NF-κB in regulating genes involved in inflammation and cell survival, understanding its activation in the context of AMD is crucial. Dysregulated NF-κB activation can lead to chronic inflammation, oxidative damage, and cell death, all of which contribute to retinal degeneration in AMD [2, 3].

This study aims to delineate the roles of these cytokines in activating the NF-κB pathway in RPE and photoreceptor cells and to evaluate how this activation influences cellular processes and contributes to AMD pathogenesis. By investigating these molecular mechanisms, the study seeks to identify potential therapeutic targets within the NF-κB pathway that could mitigate the deleterious effects of inflammation and oxidative stress in AMD [<u>4</u>. <u>5</u>].

The importance of this study lies in its potential to advance the understanding of the molecular underpinnings of AMD. By elucidating the specific contributions of cytokine-induced NF-κB activation, we can inform the development of targeted therapies aimed at preventing or slowing the progression of AMD, ultimately improving outcomes for patients affected by this debilitating disease [6, 7].

## Methods

### Aim of the Study

This study investigates the role of pro-inflammatory cytokines—Tumor Necrosis Factor-alpha (TNF-α), Interleukin-6 (IL-6), Interleukin-1 beta (IL-1β), Interleukin-8 (IL-8), Interferon-gamma (IFN-γ), Interleukin-17 (IL-17), Interleukin-12 (IL-12), Interleukin-18 (IL-18), Interleukin-33 (IL-33), and Interleukin-25 (IL-25)—in activating the NF-κB signaling pathway in retinal pigment epithelium (RPE) and photoreceptor cells. The aim is to understand the molecular mechanisms involved in this activation and to explore how it influences cellular processes, including inflammation, oxidative stress, apoptosis, and extracellular matrix remodeling, ultimately contributing to the pathogenesis of Age-related Macular Degeneration (AMD).

### Research Question

How do the cytokines TNF-α, IL-6, IL-1β, IL-8, IFN-γ, IL-17, IL-12, IL-18, IL-33, and IL-25 activate the NF-κB signaling pathway in retinal pigment epithelium and photoreceptor cells, and what molecular mechanisms drive this activation to contribute to cellular processes that lead to Age-related Macular Degeneration (AMD)?

### Search Focus

A comprehensive literature search was conducted using multiple databases, including PUBMED, MEDLINE, and Google Scholar, along with open-access and subscription-based journals. There were no date restrictions for published articles. The search specifically focused on studies investigating the activation of NF-κB signaling in RPE and photoreceptor cells by:

- TNF-α
- IL-6
- IL-1β
- IL-8
- IFN-γ
- IL-17
- IL-12
- IL-18
- IL-33
- IL-25

The literature was screened and selected based on relevance to the study’s objectives. Literature search began in September 2021 and ended in April 2024. An in-depth investigation was conducted during this duration based on the parameters of the study as defined above. During revision, further literature was searched and referenced until October 2024. The literature search and all sections of the manuscript were checked multiple times during the months of revision (May 2024 – October 2024) to maintain the highest accuracy possible. This approach allowed for a comprehensive investigation of the role of these cytokines in NF-κB pathway activation and their contribution to AMD pathogenesis, adhering to the PRISMA (Preferred Reporting Items for Systematic Reviews and Meta-Analyses) guidelines for systematic reviews.

### Search Queries/Keywords

**1. General terms:**

"Age-related Macular Degeneration" OR "AMD"
"Retinal Pigment Epithelium" OR "RPE"
"Photoreceptor Cells"
"NF-κB signaling pathway"
**2. Cytokines:**

"TNF-α" AND "NF-κB activation"
"IL-6" AND "inflammation"
"IL-1β" AND "RPE" OR "photoreceptor cells"
"IL-8" AND "oxidative stress"
"IFN-γ" AND "apoptosis"
"IL-17" AND "NF-κB signaling"
"IL-12" AND "extracellular matrix remodeling"
"IL-18" AND "AMD"
"IL-33" AND "retinal degeneration"
"IL-25" AND "chronic inflammation"

Boolean operators (AND, OR) were applied to generate targeted queries that retrieved studies addressing these cytokines’ involvement in NF-κB activation in the context of AMD and related pathophysiological processes.

### Objectives of the Searches

- To elucidate the role of TNF-α, IL-6, IL-1β, IL-8, IFN-γ, IL-17, IL-12, IL-18, IL-33, and IL-25 in activating the NF-κB signaling pathway in RPE and photoreceptor cells.
- To identify the molecular mechanisms by which these cytokines drive NF-κB-mediated cellular processes, such as inflammation, oxidative stress, apoptosis, and extracellular matrix remodeling.
- To explore the potential contribution of these cytokine-induced processes to the pathogenesis of AMD.

### Screening and Eligibility Criteria

#### Initial Screening

Initial screening was based on article titles and abstracts to filter out irrelevant studies. Those directly examining the role of TNF-α, IL-6, IL-1β, IL-8, IFN-γ, IL-17, IL-12, IL-18, IL-33, and IL-25 in NF-κB activation within RPE and photoreceptor cells were selected for full-text review.

#### Full-Text Review

Full-text articles were reviewed for eligibility based on relevance to the study’s primary objectives and methodological rigor. Only studies with detailed analyses of the molecular mechanisms involved in cytokine-driven NF-κB activation and subsequent cellular processes related to AMD were considered.

#### Data Extraction

Data from selected studies were extracted based on the role of each cytokine in NF-κB activation and their downstream effects on inflammation, oxidative stress, apoptosis, and extracellular matrix remodeling in RPE and photoreceptor cells. Key findings related to the pathogenesis of AMD were also documented.

### Inclusion and Exclusion Criteria

#### Inclusion Criteria

- Studies focusing on the role of TNF-α, IL-6, IL-1β, IL-8, IFN-γ, IL-17, IL-12, IL-18, IL-33, and IL-25 in NF-κB activation within retinal pigment epithelium and photoreceptor cells.
- Research investigating the molecular mechanisms linking NF-κB activation to inflammation, apoptosis, oxidative stress, and extracellular matrix remodeling in the context of AMD.
- Peer-reviewed articles presenting experimental, mechanistic, or review data on the interaction between these cytokines and the NF-κB pathway.

#### Exclusion Criteria

- Studies not focused on NF-κB signaling or those unrelated to retinal biology and AMD.
- Research without detailed insights into cytokine-driven NF-κB activation mechanisms.
- Conference abstracts, unpublished studies, or articles in languages other than English.

### Rationale for Screening and Inclusion

The pro-inflammatory cytokines selected for investigation—TNF-α, IL-6, IL-1β, IL-8, IFN-γ, IL-17, IL-12, IL-18, IL-33, and IL-25—are known to play key roles in immune responses and chronic inflammation. Each of these cytokines has been implicated in activating the NF-κB signaling pathway, which is crucial in driving the inflammatory and degenerative processes underlying AMD. The rationale for selecting these cytokines is based on their established involvement in:

- **TNF-α**: Potent activator of NF-κB, playing a critical role in inflammation and apoptotic processes within the retina.
- **IL-6**: Involved in chronic inflammation and has been linked to oxidative stress in retinal tissues.
- **IL-1β**: A key driver of inflammation, known to affect RPE and photoreceptor cell function in AMD.
- **IL-8**: Promotes oxidative stress and inflammation, contributing to retinal degeneration.
- **IFN-γ**: Induces apoptosis and modulates immune responses, impacting retinal cell survival.
- **IL-17**: Involved in promoting inflammatory responses and activating NF-κB in various tissues, including the retina.
- **IL-12**: Known for its role in extracellular matrix remodeling, which affects the structural integrity of retinal tissues in AMD.
- **IL-18**: Associated with chronic inflammation and retinal degeneration.
- **IL-33**: Modulates immune responses and has been linked to neuroinflammatory conditions in the retina.
- **IL-25**: Plays a role in chronic inflammatory processes, potentially exacerbating AMD progression.

### Assessment of Article Quality and Potential Biases

A thorough assessment of article quality and the identification of potential biases were integral to maintaining the validity of the findings.

#### Quality Assessment

Studies were evaluated based on experimental design, data presentation, reproducibility, and overall scientific rigor. Priority was given to peer-reviewed research that provided robust evidence of cytokine involvement in NF-κB activation and AMD pathogenesis.

#### Bias Assessment

Efforts were made to minimize biases, including:

- **Publication Bias**: A broad search strategy was used to include studies with both positive and negative findings.
- **Selection Bias**: Articles were selected based on predefined criteria to ensure objectivity.
- **Reporting Bias**: Comprehensive data extraction was performed to avoid selective reporting of results.

### Language and Publication Restrictions

Only articles published in English were considered for inclusion. No restrictions were placed on the date of publication to ensure a broad range of studies were considered. Conference abstracts and unpublished studies were excluded to prioritize peer-reviewed research with established findings.

## Results

A total of 2171 articles were identified using database searching, and 2069 were recorded after duplicates removal. 1761 were excluded after screening of title/abstract, 174 were finally excluded, and 5 articles were excluded during data extraction. These exclusions were primarily due to factors such as non-conformity with the study focus, insufficient methodological rigor, or data that did not align with the research questions. Finally, 129 articles were included as references. PRISMA Flow Diagram is Fig 1.

**Fig 1.**
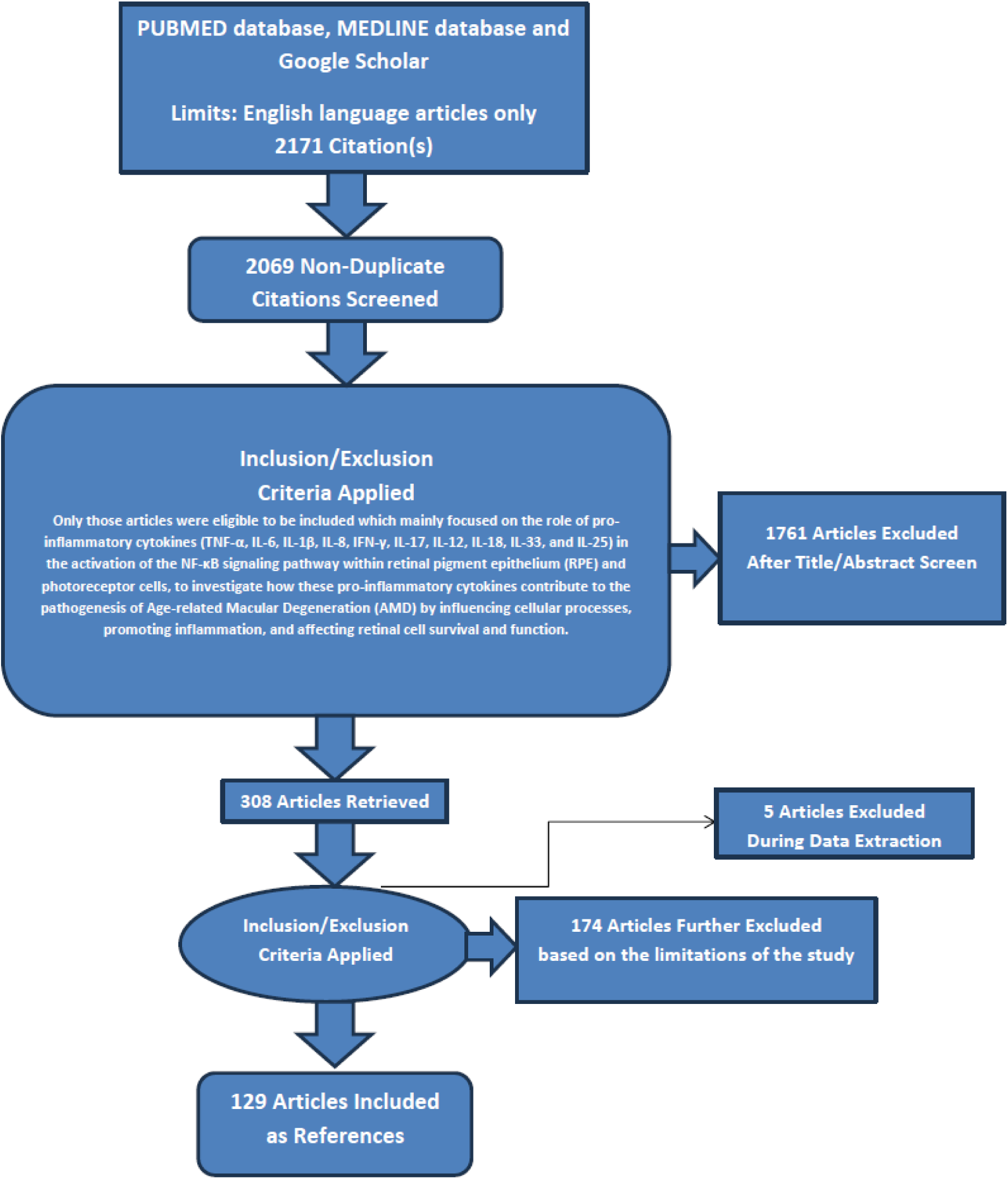
PRISMA FLOW DIAGRAM: This figure represents graphically the flow of citations in the study.

### Role of Cytokines in NF-κB Signaling Pathway in RPE and Photoreceptor Cells

#### 1. Tumor Necrosis Factor-alpha (TNF-α)

##### Molecular Mechanisms of TNF-α-Induced NF-κB Activation

Tumor Necrosis Factor-alpha (TNF-α) is a pro-inflammatory cytokine that significantly contributes to the pathogenesis of various inflammatory diseases, including Age-related Macular Degeneration (AMD). It exerts its effects primarily through the activation of the NF-κB (nuclear factor kappa-light-chain-enhancer of activated B cells) signaling pathway. This pathway plays a crucial role in regulating immune response and inflammation, making TNF-α a key player in the molecular mechanisms underlying inflammatory processes in retinal diseases [8, 11, 114].

TNF-α binds to its receptors, TNFR1 (TNF receptor 1) and TNFR2 (TNF receptor 2), on the surface of retinal pigment epithelium (RPE) and photoreceptor cells. TNFR1 predominantly mediates the activation of the NF-κB pathway. Upon binding of TNF-α, TNFR1 recruits the adaptor protein TRADD (TNF receptor-associated death domain), which acts as a scaffold for the assembly of additional signaling molecules. This recruitment is critical for the subsequent steps in the signaling cascade that lead to NF-κB activation [9, 12].

The TRADD adaptor protein then recruits RIP1 (receptor-interacting protein 1) and TRAF2 (TNF receptor-associated factor 2), forming a complex that activates the IKK (IκB kinase) complex. The IKK complex, consisting of IKKα, IKKβ, and NEMO (NF-κB essential modulator), phosphorylates IκB (inhibitor of NF-κB), marking it for ubiquitination and proteasomal degradation. This degradation releases NF-κB dimers, typically p65 and p50, which translocate into the nucleus. Once in the nucleus, NF-κB dimers bind to specific DNA sequences to promote the transcription of target genes involved in inflammation, cell survival, and immune responses [10,13,115].

##### TNF-α-Induced NF-κB Activation and Cellular Processes

TNF-α-induced NF-κB activation in retinal pigment epithelium (RPE) and photoreceptor cells influences several cellular processes that contribute to the pathogenesis of Age-related Macular Degeneration (AMD). This activation triggers the transcription of pro-inflammatory cytokines, such as IL-1β, IL-6, and IL-8, along with chemokines like MCP-1 and adhesion molecules like ICAM-1. The result is an amplification of local inflammation and the recruitment of immune cells to the retina, exacerbating tissue damage [14, 17, 124].

NF-κB signaling also increases oxidative stress by upregulating enzymes such as inducible nitric oxide synthase (iNOS) and cyclooxygenase-2 (COX-2), leading to enhanced production of reactive oxygen species (ROS).

Elevated ROS levels inflict damage on RPE and photoreceptor cells, contributing to the progression of AMD. Additionally, NF-κB promotes the expression of anti-apoptotic genes like Bcl-2 and Bcl-xL, as well as cell survival factors, which can offer protection under stress conditions. However, chronic activation of NF-κB may cause cellular dysfunction and death, furthering retinal degeneration [15, 17].

In the context of wet AMD, NF-κB activation heightens the expression of vascular endothelial growth factor (VEGF) and other angiogenic factors, encouraging the growth of abnormal blood vessels beneath the retina. These vessels are fragile and prone to leaking, leading to vision loss. Furthermore, NF-κB modulates the immune response in the retina by regulating the expression of MHC molecules and co-stimulatory factors. Dysregulated immune responses can result in chronic inflammation and subsequent tissue damage, underpinning the complex interactions of mechanisms driving AMD pathology [16, 17].

##### Contribution to AMD Pathogenesis

TNF-α-induced NF-κB activation plays a significant role in the pathogenesis of Age-related Macular Degeneration (AMD) by maintaining a chronic inflammatory state in the retina. This persistent inflammation progressively damages retinal pigment epithelium (RPE) and photoreceptor cells, driving the development of both dry and wet forms of AMD. Inflammation is a central factor in the disease’s progression, as it disrupts cellular functions and promotes tissue damage [18, 111, 118].

RPE cells are essential for maintaining retinal homeostasis, but NF-κB-mediated inflammation and oxidative stress compromise their function. This dysfunction leads to the death of photoreceptor cells and subsequent vision loss. The combined effects of increased oxidative stress and inflammatory cytokines result in direct damage to photoreceptor cells, contributing to their degeneration and the atrophy characteristic of AMD. The death of these cells exacerbates the visual impairment associated with the disease [19, 112, 117].

In wet AMD, NF-κB-driven production of vascular endothelial growth factor (VEGF) results in pathological neovascularization. This abnormal growth of blood vessels can cause retinal detachment and hemorrhages, leading to rapid vision deterioration. Additionally, NF-κB activation is implicated in the formation and accumulation of drusen, extracellular deposits that are hallmarks of AMD. The presence of drusen further exacerbates inflammation and RPE dysfunction, creating a cycle of damage that advances the disease’s progression [20, 113].

#### 2. Interleukin-6 (IL-6)

##### Molecular Mechanisms of IL-6-Induced NF-κB Activation

Interleukin-6 (IL-6) is a multifunctional cytokine that plays a critical role in inflammation, immune responses, and cellular survival. In the context of Age-related Macular Degeneration (AMD), IL-6 can activate the NF-κB (nuclear factor kappa-light-chain-enhancer of activated B cells) signaling pathway in retinal pigment epithelium (RPE) and photoreceptor cells, contributing to the disease’s pathogenesis. This activation underscores the significance of IL-6 in mediating inflammatory processes that exacerbate retinal degeneration [21, 111].

IL-6 signals through a receptor complex consisting of the IL-6 receptor (IL-6R) and the signal transducer glycoprotein 130 (gp130). Upon binding of IL-6 to IL-6R, gp130 undergoes dimerization, initiating downstream signaling cascades. The IL-6/IL-6R/gp130 complex activates Janus kinases (JAKs), leading to the phosphorylation of STAT3 (signal transducer and activator of transcription 3). This pathway is crucial for the transcriptional regulation of various genes involved in inflammation and cellular survival, thereby playing a crucial role in the cellular responses associated with AMD [22, 113].

IL-6 signaling can indirectly activate the NF-κB pathway through several mechanisms. The activation of the IκB kinase (IKK) complex results in the phosphorylation and degradation of IκB proteins, releasing NF-κB dimers.

Additionally, activated STAT3 can interact with NF-κB components, enhancing NF-κB’s transcriptional activity on target genes. In certain contexts, IL-6 may directly activate the NF-κB pathway via other signaling intermediates, such as phosphoinositide 3-kinase (PI3K) and AKT, which contribute to the activation of IKK. These interactions highlight the complex cross-talk between IL-6 signaling and NF-κB activation, emphasizing their collaborative role in the pathogenesis of AMD [23, 114].

##### IL-6-Induced NF-κB Activation and Cellular Processes

IL-6-induced NF-κB activation in retinal pigment epithelium (RPE) and photoreceptor cells affects several cellular processes contributing to the pathogenesis of Age-related Macular Degeneration (AMD). One key outcome of NF-κB activation is the upregulation of pro-inflammatory cytokines, such as IL-1β and TNF-α, and chemokines like MCP-1 and IL-8. This cytokine and chemokine production establishes a feedback loop that sustains and amplifies inflammation within the retina, exacerbating the degenerative processes associated with AMD [24, 115, 125].

NF-κB activation also enhances the expression of enzymes that generate reactive oxygen species (ROS), including inducible nitric oxide synthase (iNOS) and NADPH oxidase. The increased oxidative stress resulting from elevated ROS levels damages RPE and photoreceptor cells, further contributing to the progression of AMD. The chronic oxidative environment within the retina is a significant factor in the pathophysiology of this disease, leading to cellular dysfunction and death [25, 119].

IL-6-induced NF-κB activation promotes angiogenesis by upregulating the expression of vascular endothelial growth factor (VEGF), leading to the formation of abnormal blood vessels characteristic of wet AMD. These new vessels are prone to leaking fluid and blood, causing rapid vision loss. Additionally, NF-κB regulates genes involved in cell survival, such as Bcl-2 and Bcl-xL, and apoptosis. While acute NF-κB activation may promote cell survival, chronic activation can lead to cellular dysfunction and apoptosis, contributing to retinal degeneration. NF-κB also modulates immune responses in the retina by regulating MHC molecules and co-stimulatory signals, with dysregulated immune responses leading to chronic inflammation and tissue damage [26, 118].

##### Contribution to AMD Pathogenesis

IL-6-induced NF-κB activation plays a significant role in the pathogenesis of Age-related Macular Degeneration (AMD) through various mechanisms. Chronic activation of NF-κB maintains a state of persistent inflammation in the retina, driving progressive damage to retinal pigment epithelium (RPE) and photoreceptor cells. This chronic inflammation is a critical factor in both dry and wet AMD, promoting cellular injury and disease progression [27,117].

Disruption of RPE function is another crucial aspect of IL-6-induced NF-κB activation. RPE cells are essential for maintaining retinal homeostasis, and their dysfunction, triggered by inflammation and oxidative stress, leads to photoreceptor cell death and subsequent vision loss. Additionally, inflammatory cytokines and increased oxidative stress directly damage photoreceptor cells, accelerating their atrophy and degeneration, a hallmark of AMD [28, 112].

In wet AMD, NF-κB-driven production of vascular endothelial growth factor (VEGF) promotes pathological neovascularization, resulting in retinal detachment and hemorrhages that rapidly deteriorate vision.

Furthermore, NF-κB activation may facilitate the formation and accumulation of drusen, extracellular deposits that are characteristic of AMD. This accumulation exacerbates inflammation and RPE dysfunction, further contributing to the progression of the disease [29, 30].

#### 3. Interleukin-1 beta (IL-1β)

##### Molecular Mechanisms of IL-1β-Induced NF-κB Activation

Interleukin-1 beta (IL-1β) is a potent pro-inflammatory cytokine that plays a critical role in the inflammatory and immune responses associated with Age-related Macular Degeneration (AMD). In AMD, IL-1β activates the NF-κB (nuclear factor kappa-light-chain-enhancer of activated B cells) signaling pathway within retinal pigment epithelium (RPE) and photoreceptor cells. This pathway is crucial for regulating the expression of genes involved in inflammation and cell survival [31, 113].

The activation process begins when IL-1β binds to the IL-1 receptor type I (IL-1RI) on the surface of RPE and photoreceptor cells. This receptor-ligand interaction necessitates the involvement of the IL-1 receptor accessory protein (IL-1RAcP) to facilitate signal transduction. Following this binding, the IL-1RI/IL-1RAcP complex recruits the adaptor protein MyD88 (myeloid differentiation primary response 88), which is indispensable for the downstream signaling cascade [32, 114].

Once MyD88 is recruited, it activates members of the IL-1 receptor-associated kinase (IRAK) family, specifically IRAK1 and IRAK4. These activated IRAKs then interact with TNF receptor-associated factor 6 (TRAF6), leading to the activation of TGF-β-activated kinase 1 (TAK1). TAK1 subsequently phosphorylates and activates the IκB kinase (IKK) complex, which consists of IKKα, IKKβ, and NEMO (NF-κB essential modulator). The IKK complex phosphorylates IκB proteins, marking them for ubiquitination and degradation, which releases NF-κB dimers, typically p65 and p50. These freed NF-κB dimers translocate to the nucleus, where they bind to specific DNA sequences and promote the transcription of genes that drive inflammation, cell survival, and immune responses [33, 115].

##### IL-1β-Induced NF-κB Activation and Cellular Processes

IL-1β-induced NF-κB activation in retinal pigment epithelium (RPE) and photoreceptor cells significantly impacts several cellular processes contributing to the pathogenesis of Age-related Macular Degeneration (AMD). One of the critical effects is the transcription of pro-inflammatory cytokines such as IL-6 and TNF-α, and chemokines like MCP-1 and IL-8. This transcriptional activity creates a feedback loop that sustains and amplifies inflammation within the retina, exacerbating the inflammatory environment characteristic of AMD [34, 111].

Another crucial aspect is the role of NF-κB activation in enhancing oxidative stress. The expression of enzymes that generate reactive oxygen species (ROS), such as inducible nitric oxide synthase (iNOS) and NADPH oxidase, is upregulated, leading to increased oxidative stress. The resulting ROS damage RPE and photoreceptor cells, thereby contributing to the progression of AMD. Oxidative stress not only damages cellular components but also perpetuates the inflammatory cycle, creating a deleterious environment within the retina [35, 119, 126].

IL-1β-induced NF-κB activation also promotes angiogenesis, particularly through the upregulation of vascular endothelial growth factor (VEGF). In wet AMD, this leads to the formation of abnormal blood vessels that can leak fluid and blood, causing rapid vision loss. Additionally, NF-κB influences cell survival and apoptosis by regulating genes such as Bcl-2 and Bcl-xL. While acute activation of NF-κB can promote cell survival, chronic activation may result in cellular dysfunction and death, contributing to retinal degeneration. Furthermore, NF-κB modulates immune responses by regulating the expression of MHC molecules and co-stimulatory signals, and dysregulation of these immune responses can lead to chronic inflammation and tissue damage in the retina [<u>36</u>,

118].

##### Contribution to AMD Pathogenesis

IL-1β-induced NF-κB activation plays a significant role in the pathogenesis of Age-related Macular Degeneration (AMD) through several mechanisms. Chronic inflammation in the retina is sustained by this activation, leading to progressive damage to retinal pigment epithelium (RPE) and photoreceptor cells. This inflammation is a key driver of both the dry and wet forms of AMD, perpetuating a cycle of cellular damage and dysfunction [37, 117].

RPE cells, essential for maintaining retinal homeostasis, are particularly affected by IL-1β-induced NF-κB activation. The resulting inflammation and oxidative stress disrupt RPE function, leading to photoreceptor cell death and subsequent vision loss. The direct damage to photoreceptor cells from inflammatory cytokines and oxidative stress contributes to their degeneration, which is a hallmark of AMD progression [38, 112].

In wet AMD, NF-κB-driven production of vascular endothelial growth factor (VEGF) leads to pathological neovascularization, causing retinal detachment and hemorrhages, which result in rapid vision deterioration. Additionally, NF-κB activation may contribute to the formation and accumulation of drusen, extracellular deposits that are characteristic of AMD. The accumulation of drusen exacerbates inflammation and RPE dysfunction, further contributing to the disease’s progression [39, 40].

#### 4. Interleukin-8 (IL-8)

##### Molecular Mechanisms of IL-8-Induced NF-κB Activation

Interleukin-8 (IL-8), also known as CXCL8, is a pro-inflammatory chemokine that plays a crucial role in the recruitment and activation of neutrophils. In the context of Age-related Macular Degeneration (AMD), IL-8 can activate the NF-κB signaling pathway in retinal pigment epithelium (RPE) and photoreceptor cells, contributing significantly to the disease’s pathogenesis [41, 113].

IL-8 exerts its effects through binding to its receptors, CXCR1 and CXCR2, which are G protein-coupled receptors expressed on RPE and photoreceptor cells. Upon binding, IL-8 activates G proteins, initiating downstream signaling pathways such as PI3K/Akt and Ras/Raf/MEK/ERK. These pathways subsequently lead to the activation of the IκB kinase (IKK) complex, consisting of IKKα, IKKβ, and NEMO (NF-κB essential modulator) [42, 114].

The activation of the IKK complex results in the phosphorylation of IκB proteins, marking them for ubiquitination and proteasomal degradation. This degradation releases NF-κB dimers, typically composed of p65 and p50, from their inhibitory IκB proteins. The freed NF-κB dimers then translocate to the nucleus, where they bind to specific DNA sequences, initiating the transcription of target genes involved in inflammation, cell survival, and immune responses [43, 115].

##### IL-8-Induced NF-κB Activation and Cellular Processes

IL-8-induced NF-κB activation in retinal pigment epithelium (RPE) and photoreceptor cells impacts various cellular processes critical to the pathogenesis of Age-related Macular Degeneration (AMD). Activation of NF-κB leads to the upregulation of pro-inflammatory cytokines, such as TNF-α and IL-1β, and chemokines like MCP-1. This promotes local inflammation and recruits additional immune cells to the retina, exacerbating the inflammatory environment [44, 119].

Enhanced NF-κB activation also contributes to increased oxidative stress by upregulating the expression of enzymes responsible for generating reactive oxygen species (ROS), including NADPH oxidase and inducible nitric oxide synthase (iNOS). The resultant oxidative damage to RPE and photoreceptor cells accelerates AMD progression. Concurrently, IL-8 functions as a pro-angiogenic factor, further amplified by NF-κB activation, which increases the expression of vascular endothelial growth factor (VEGF) and other angiogenic factors. This promotes the formation of abnormal blood vessels in wet AMD, leading to leakage and rapid vision loss [45, 111].

The role of NF-κB in regulating cell survival and apoptosis is also significant. It modulates the expression of genes involved in cell survival, such as Bcl-2 and Bcl-xL, as well as apoptotic pathways. Acute NF-κB activation may enhance cell survival; however, chronic activation can result in cellular dysfunction and death, contributing to retinal degeneration. Additionally, NF-κB influences immune modulation by regulating the expression of MHC molecules and co-stimulatory signals, which can impact immune responses in the retina and potentially lead to chronic inflammation and tissue damage [46, 118].

##### Contribution to AMD Pathogenesis

IL-8-induced NF-κB activation plays a significant role in the pathogenesis of Age-related Macular Degeneration (AMD) through various mechanisms. Chronic activation of NF-κB leads to a persistent inflammatory state in the retina, which progressively damages retinal pigment epithelium (RPE) and photoreceptor cells. This chronic inflammation is a critical factor driving both dry and wet forms of AMD [47, 112].

The function of RPE cells, essential for maintaining retinal homeostasis, is disrupted by IL-8-induced NF-κB activation. This disruption results from both inflammatory processes and oxidative stress, ultimately causing photoreceptor cell death and contributing to vision loss. Direct damage to photoreceptor cells from inflammatory cytokines and oxidative stress accelerates their atrophy, further exacerbating the progression of AMD [48, 117].

In wet AMD, NF-κB-driven production of vascular endothelial growth factor (VEGF) promotes pathological neovascularization, leading to retinal detachment and hemorrhages that significantly impair vision. Additionally, NF-κB activation may facilitate the formation and accumulation of drusen, extracellular deposits characteristic of AMD. The presence of drusen further aggravates inflammation and RPE dysfunction, compounding the disease’s progression [49, 50].

#### 5. Interferon-gamma (IFN-γ)

##### Molecular Mechanisms of IFN-γ-Induced NF-κB Activation

Interferon-gamma (IFN-γ) plays a pivotal role in both innate and adaptive immunity due to its potent pro-inflammatory properties. In Age-related Macular Degeneration (AMD), IFN-γ significantly impacts the activation of the NF-κB (nuclear factor kappa-light-chain-enhancer of activated B cells) signaling pathway within retinal pigment epithelium (RPE) and photoreceptor cells. This activation is a critical component in the progression of the disease [51, 113].

Upon binding to its receptor, which consists of IFN-γR1 and IFN-γR2 subunits, IFN-γ triggers the activation of Janus kinases (JAK1 and JAK2). These kinases phosphorylate the receptor, creating docking sites for STAT1 (signal transducer and activator of transcription 1). The phosphorylated STAT1 then dimerizes and moves to the nucleus, where it binds to gamma-activated sequences (GAS) in the promoters of IFN-γ-responsive genes, initiating the transcription of various target genes [52, 114].

The JAK-STAT pathway interacts with the NF-κB pathway at multiple levels. STAT1 activation upregulates genes encoding NF-κB subunits and regulatory proteins. Additionally, IFN-γ signaling can activate the IκB kinase (IKK) complex through intermediary proteins such as TRAF (TNF receptor-associated factor) and TAK1 (TGF-β-activated kinase 1). The activated IKK complex phosphorylates IκB proteins, leading to their ubiquitination and subsequent degradation. This process releases NF-κB dimers, such as p65 and p50, which then translocate to the nucleus to bind DNA sequences and drive the transcription of genes involved in inflammation, cell survival, and immune responses [53,115].

##### IFN-γ-Induced NF-κB Activation and Cellular Processes

IFN-γ-induced activation of the NF-κB signaling pathway in retinal pigment epithelium (RPE) and photoreceptor cells influences several cellular processes integral to Age-related Macular Degeneration (AMD). One major consequence is the enhanced production of inflammatory cytokines such as TNF-α, IL-1β, and IL-6, along with chemokines like MCP-1. This upregulation intensifies local inflammation and attracts additional immune cells to the retina, exacerbating the inflammatory response [54, 119].

Increased oxidative stress is another outcome of NF-κB activation. The pathway promotes the expression of enzymes responsible for generating reactive oxygen species (ROS), including NADPH oxidase and inducible nitric oxide synthase (iNOS). Elevated levels of ROS inflict damage on RPE and photoreceptor cells, accelerating the progression of AMD. This oxidative damage contributes significantly to retinal cell dysfunction and degeneration [55, 118, 127].

The modulation of immune responses by IFN-γ further complicates AMD pathology. By upregulating MHC class II molecules and other co-stimulatory factors, IFN-γ enhances antigen presentation and immune cell activation, which can lead to persistent inflammation in the retina. Additionally, NF-κB influences cell survival and apoptosis by regulating genes such as Bcl-2 and Bcl-xL. While acute NF-κB activation may support cell survival, prolonged activation often leads to cellular dysfunction and death. Although not a primary angiogenic factor, IFN-γ’s effects on inflammatory cytokines and NF-κB can indirectly foster angiogenesis, contributing to the development of abnormal blood vessels in wet AMD [56, 117].

##### Contribution to AMD Pathogenesis

IFN-γ-induced activation of the NF-κB signaling pathway plays a significant role in the pathogenesis of Age-related Macular Degeneration (AMD) through various mechanisms. Chronic NF-κB activation establishes a persistent inflammatory environment in the retina, which leads to ongoing damage of retinal pigment epithelium (RPE) and photoreceptor cells. This chronic inflammation is a major factor driving both dry and wet forms of AMD [57, 112].

Disruption of RPE function is another critical consequence of NF-κB activation. RPE cells are essential for maintaining retinal health, but their function is compromised by inflammation and oxidative stress induced by IFN-γ. This dysfunction ultimately results in the death of photoreceptor cells and subsequent vision loss [58, 111,128].

Photoreceptor degeneration is directly impacted by inflammatory cytokines and oxidative stress, which lead to the atrophy and loss of these cells. In the case of wet AMD, NF-κB activation promotes the production of vascular endothelial growth factor (VEGF), driving pathological neovascularization that causes retinal detachment and hemorrhage, leading to rapid vision deterioration. Additionally, NF-κB activation may facilitate the formation and accumulation of drusen, extracellular deposits that are characteristic of AMD and exacerbate inflammation and RPE dysfunction [59, 60].

#### 6. Interleukin-17 (IL-17)

##### Molecular Mechanisms of IL-17-Induced NF-κB Activation

Interleukin-17 (IL-17), a pro-inflammatory cytokine predominantly produced by Th17 cells, is crucial in the progression of autoimmune and inflammatory conditions. In Age-related Macular Degeneration (AMD), IL-17 has been implicated in disease pathogenesis through its activation of the NF-κB (nuclear factor kappa-light-chain-enhancer of activated B cells) signaling pathway within retinal pigment epithelium (RPE) and photoreceptor cells. The interaction between IL-17 and the NF-κB pathway highlights a critical mechanism in AMD pathology, emphasizing the need to understand these molecular interactions [61, 114].

The activation of IL-17 signaling begins with the cytokine binding to its receptor complex, which includes the IL-17RA and IL-17RC subunits present on the surface of RPE and photoreceptor cells. This interaction initiates a cascade of intracellular events, starting with the recruitment of adaptor proteins such as Act1 (nuclear factor NF-kappa-B activator 1). Act1 is essential for propagating the signal downstream, leading to the recruitment and activation of TRAF6 (TNF receptor-associated factor 6) [62, 113].

TRAF6, an E3 ubiquitin ligase, subsequently activates the IκB kinase (IKK) complex, which consists of IKKα, IKKβ, and NEMO (NF-κB essential modulator). The activated IKK complex phosphorylates IκB proteins, marking them for ubiquitination and proteasomal degradation. This degradation releases NF-κB dimers, typically p65 and p50, which then translocate to the nucleus. In the nucleus, NF-κB dimers bind to specific DNA sequences, driving the transcription of genes involved in inflammation, cell survival, and immune responses [63, 115, 129].

##### IL-17-Induced NF-κB Activation and Cellular Processes

IL-17-induced NF-κB activation in retinal pigment epithelium (RPE) and photoreceptor cells influences several cellular processes that contribute to Age-related Macular Degeneration (AMD). The activation of NF-κB results in the upregulation of pro-inflammatory cytokines such as TNF-α, IL-1β, and IL-6, as well as chemokines including IL-8 and MCP-1. This upregulation amplifies local inflammation and recruits additional immune cells to the retina, exacerbating the inflammatory response and potentially accelerating disease progression [64, 119].

NF-κB activation also enhances the expression of enzymes involved in oxidative stress, such as NADPH oxidase and inducible nitric oxide synthase (iNOS). The increased production of reactive oxygen species (ROS) leads to oxidative damage in RPE and photoreceptor cells, further contributing to the advancement of AMD. This oxidative stress not only damages cellular components but also disrupts normal cellular functions, exacerbating retinal degeneration [65, 111].

In addition to inflammatory and oxidative responses, IL-17-induced NF-κB activation modulates immune responses by increasing the expression of MHC class II molecules and other co-stimulatory molecules. This enhances antigen presentation and immune cell activation, potentially leading to sustained inflammation within the retina. NF-κB also regulates genes involved in cell survival and apoptosis, with acute activation supporting cell survival and chronic activation promoting cellular dysfunction and death. Although IL-17 itself is not a primary angiogenic factor, its role in inducing pro-inflammatory cytokines and activating NF-κB can indirectly foster angiogenesis, contributing to the development of abnormal blood vessels in wet AMD [66, 118].

##### Contribution to AMD Pathogenesis

IL-17-induced NF-κB activation plays a significant role in the pathogenesis of Age-related Macular Degeneration (AMD) through various mechanisms. Chronic activation of NF-κB sustains a prolonged inflammatory state within the retina, which progressively damages retinal pigment epithelium (RPE) and photoreceptor cells. This persistent inflammation is a crucial factor in the development of both dry and wet forms of AMD, driving disease progression and retinal dysfunction [67, 112].

The function of RPE cells, essential for maintaining retinal homeostasis, is disrupted by IL-17-induced NF-κB activation. Inflammation and oxidative stress from this activation impair RPE function, leading to the death of photoreceptor cells and subsequent vision loss. Additionally, inflammatory cytokines and oxidative stress directly damage photoreceptor cells, contributing to their atrophy and loss, which further accelerates AMD [68, 117].

In wet AMD, NF-κB-driven production of vascular endothelial growth factor (VEGF) induces pathological neovascularization. This process results in retinal detachment and hemorrhages, causing rapid deterioration in vision. NF-κB activation may also influence the formation and accumulation of drusen, extracellular deposits characteristic of AMD, which exacerbate both inflammation and RPE dysfunction, compounding the progression of the disease [69, 70].

#### 7. Interleukin-12 (IL-12)

##### Molecular Mechanisms of IL-12-Induced NF-κB Activation

Interleukin-12 (IL-12), a pro-inflammatory cytokine predominantly produced by dendritic cells, macrophages, and B cells, is essential in orchestrating immune responses. It facilitates the differentiation of naive T cells into Th1 cells and augments the cytotoxic functions of natural killer (NK) cells and CD8+ T cells. In the pathology of Age-related Macular Degeneration (AMD), IL-12 plays a significant role by activating the NF-κB (nuclear factor kappa-light-chain-enhancer of activated B cells) signaling pathway within retinal pigment epithelium (RPE) and photoreceptor cells, thereby contributing to disease progression [71, 111].

The biological effects of IL-12 are mediated through its interaction with a receptor complex composed of IL-12Rβ1 and IL-12Rβ2. These receptors are present on the surface of various immune cells and also on RPE and photoreceptor cells. IL-12 binding to this receptor complex activates the JAK-STAT signaling pathway, leading to the phosphorylation and activation of JAK2 and TYK2. These kinases subsequently phosphorylate STAT4, which is critical for the next steps in the signaling cascade [72, 114].

Once phosphorylated, STAT4 dimerizes and translocates to the nucleus, where it binds to specific DNA sequences in the promoters of IL-12-responsive genes, initiating their transcription. The JAK-STAT pathway’s activation intersects with the NF-κB pathway through several mechanisms. Notably, STAT4 enhances the expression of NF-κB subunits and regulatory proteins, while IL-12 signaling can activate the IκB kinase (IKK) complex. This complex phosphorylates IκB proteins, leading to their degradation and the subsequent release of NF-κB dimers (such as p65 and p50). These free NF-κB dimers then migrate to the nucleus, where they initiate the transcription of genes involved in inflammation, cell survival, and immune responses [73, 115].

##### IL-12-Induced NF-κB Activation and Cellular Processes

IL-12-induced activation of the NF-κB pathway in retinal pigment epithelium (RPE) and photoreceptor cells influences various cellular processes that are integral to the development of Age-related Macular Degeneration (AMD). Activation of NF-κB leads to the increased production of pro-inflammatory cytokines, such as TNF-α, IL-1β, and IL-6, as well as chemokines like IL-8 and MCP-1. This upregulation amplifies local inflammation within the retina and attracts additional immune cells, exacerbating the inflammatory response [74, 119].

Increased NF-κB activity also enhances oxidative stress by promoting the expression of enzymes that produce reactive oxygen species (ROS), including NADPH oxidase and inducible nitric oxide synthase (iNOS). Elevated levels of oxidative stress result in damage to RPE and photoreceptor cells, further advancing the progression of AMD. The chronic oxidative damage contributes to cellular dysfunction and loss, intensifying the disease’s impact [75, 118].

IL-12’s modulation of immune responses involves the upregulation of MHC class II molecules and other co-stimulatory factors, which enhances antigen presentation and immune cell activation. This immune activation can result in prolonged inflammation within the retina. Additionally, NF-κB influences cell survival and apoptosis by regulating genes such as Bcl-2 and Bcl-xL. While acute NF-κB activation supports cell survival, persistent activation can lead to cellular dysfunction and death, contributing to retinal degeneration. Although IL-12 itself is not a direct angiogenic factor, its role in inducing inflammatory cytokines and activating NF-κB can indirectly promote angiogenesis, leading to the formation of abnormal blood vessels observed in wet AMD [76, 113].

##### Contribution to AMD Pathogenesis

IL-12-induced NF-κB activation plays a significant role in the pathogenesis of Age-related Macular Degeneration (AMD) through multiple mechanisms. Chronic activation of NF-κB sustains an inflammatory environment within the retina, causing progressive damage to retinal pigment epithelium (RPE) and photoreceptor cells. This ongoing inflammation is a fundamental factor in the progression of both dry and wet forms of AMD [77, 112, 123].

The disruption of RPE function, essential for maintaining retinal homeostasis, results from IL-12-induced NF-κB activation. This disruption occurs through mechanisms of inflammation and oxidative stress, leading to photoreceptor cell death and subsequent vision loss. In parallel, inflammatory cytokines and oxidative stress directly impair photoreceptor cells, accelerating their atrophy and loss, which is a characteristic feature of AMD [78, 117].

In wet AMD, the activation of NF-κB drives the production of vascular endothelial growth factor (VEGF), leading to pathological neovascularization. This process results in retinal detachment and hemorrhage, significantly impairing vision. Additionally, NF-κB activation may contribute to drusen formation—extracellular deposits that are a hallmark of AMD. The accumulation of drusen exacerbates both inflammation and RPE dysfunction, further advancing the disease [79, 80].

#### 8. Interleukin-18 (IL-18)

##### Molecular Mechanisms of IL-18-Induced NF-κB Activation

Interleukin-18 (IL-18), a pro-inflammatory cytokine from the IL-1 family, is produced by macrophages and various other cell types and plays a crucial role in both innate and adaptive immunity. It is particularly involved in stimulating interferon-gamma (IFN-γ) production by T cells and natural killer (NK) cells. In Age-related Macular Degeneration (AMD), IL-18 is known to activate the NF-κB (nuclear factor kappa-light-chain-enhancer of activated B cells) signaling pathway within retinal pigment epithelium (RPE) and photoreceptor cells, thereby contributing to the progression of the disease [89, 90].

IL-18 exerts its effects by binding to a receptor complex composed of IL-18Rα and IL-18Rβ, which are present on the surface of immune cells as well as RPE and photoreceptor cells. The binding of IL-18 to this receptor complex is crucial for initiating downstream signaling events. This interaction leads to the recruitment of the adaptor protein MyD88 (myeloid differentiation primary response 88), which is essential for the subsequent activation of the signaling cascade [81, 111].

Upon MyD88 recruitment, IL-1 receptor-associated kinases (IRAKs), specifically IRAK4 and IRAK1, are recruited and activated. These kinases then interact with TRAF6 (TNF receptor-associated factor 6), which functions as an E3 ubiquitin ligase. The activation of TRAF6 promotes the ubiquitination and activation of TAK1 (TGF-β-activated kinase 1). TAK1, in turn, activates the IκB kinase (IKK) complex consisting of IKKα, IKKβ, and NEMO (NF-κB essential modulator). The activated IKK complex phosphorylates IκB proteins, leading to their ubiquitination and degradation by the proteasome. This degradation releases NF-κB dimers, such as p65 and p50, which then translocate to the nucleus to bind DNA sequences and initiate the transcription of genes involved in inflammation, cell survival, and immune responses [82, 113].

##### IL-18-Induced NF-κB Activation and Cellular Processes

IL-18-induced activation of NF-κB in retinal pigment epithelium (RPE) and photoreceptor cells impacts various cellular processes that are central to the pathogenesis of Age-related Macular Degeneration (AMD). This activation results in the upregulation of pro-inflammatory cytokines such as TNF-α, IL-1β, and IL-6, as well as chemokines like IL-8 and MCP-1. These factors amplify local inflammation and attract additional immune cells to the retina, exacerbating inflammatory responses and contributing to disease progression [83, 114].

The activation of NF-κB also influences oxidative stress within the retina. It enhances the expression of enzymes responsible for generating reactive oxygen species (ROS), including NADPH oxidase and inducible nitric oxide synthase (iNOS). Elevated levels of ROS inflict oxidative damage on RPE and photoreceptor cells, further accelerating the degenerative processes associated with AMD [84, 115].

Additionally, IL-18 modulates immune responses by increasing the expression of MHC class II molecules and co-stimulatory molecules, which enhances antigen presentation and immune cell activation. This heightened immune response can lead to persistent inflammation within the retina. NF-κB activation also plays a role in regulating cell survival and apoptosis. While acute NF-κB activation can promote cell survival by upregulating anti-apoptotic factors such as Bcl-2 and Bcl-xL, chronic activation may induce cellular dysfunction and death, contributing to retinal degeneration. Although not primarily an angiogenic factor, IL-18’s role in inducing other pro-inflammatory cytokines and activating NF-κB can indirectly foster angiogenesis, which is associated with the formation of abnormal blood vessels in wet AMD [85, 118].

##### Contribution to AMD Pathogenesis

IL-18-induced activation of NF-κB plays a significant role in the pathogenesis of Age-related Macular Degeneration (AMD) through various mechanisms. Persistent NF-κB activation leads to chronic inflammation within the retina, which progressively damages retinal pigment epithelium (RPE) and photoreceptor cells. This ongoing inflammatory state is a crucial factor driving both dry and wet forms of AMD, contributing to the deterioration of retinal health [86, 117].

RPE cells are essential for maintaining retinal homeostasis, and their dysfunction is a critical aspect of AMD. The inflammatory and oxidative stress induced by IL-18-driven NF-κB activation disrupts RPE function, resulting in the death of photoreceptor cells and subsequent vision loss. The damage to RPE cells impairs their ability to support photoreceptors, exacerbating retinal degeneration [87, 112].

The impact of IL-18-induced NF-κB activation extends to photoreceptor degeneration and neovascularization. Inflammatory cytokines and increased oxidative stress cause direct harm to photoreceptor cells, leading to their atrophy and loss. In the case of wet AMD, NF-κB-mediated production of vascular endothelial growth factor (VEGF) promotes pathological neovascularization, which can cause retinal detachment and hemorrhages, resulting in rapid vision decline. Additionally, NF-κB activation may contribute to the formation and accumulation of drusen, extracellular deposits characteristic of AMD, further aggravating inflammation and RPE dysfunction [88, 89, 122].

#### 9. Interleukin-33 (IL-33)

##### Molecular Mechanisms of IL-33-Induced NF-κB Activation

Interleukin-33 (IL-33), a cytokine from the IL-1 family, functions as an alarmin released in response to cellular damage or stress. It initiates signaling through its receptor complex, which includes ST2 (IL-1 receptor-like 1) and IL-1 receptor accessory protein (IL-1RAcP). The primary pathway activated by IL-33 involves the NF-κB signaling cascade. IL-33 first binds to the ST2 receptor and IL-1RAcP complex present on the surface of retinal pigment epithelium (RPE) and photoreceptor cells [91, 113].

Upon binding, IL-33 recruits the adaptor protein MyD88 (myeloid differentiation primary response 88) to the receptor complex. MyD88 then facilitates the recruitment and activation of IL-1 receptor-associated kinases (IRAKs), particularly IRAK1 and IRAK4. These activated IRAKs, in turn, phosphorylate and activate TRAF6 (TNF receptor-associated factor 6), an E3 ubiquitin ligase, which plays a crucial role in further signaling [92, 114].

The activation of TRAF6 leads to the activation of TAK1 (TGF-β-activated kinase 1), which subsequently phosphorylates and activates the IκB kinase (IKK) complex consisting of IKKα, IKKβ, and NEMO (NF-κB essential modulator). This activation causes phosphorylation and degradation of inhibitory IκB proteins by the proteasome, freeing NF-κB dimers, such as p65 and p50. Once released, these dimers translocate to the nucleus where they bind to specific DNA sequences, initiating transcription of genes involved in inflammation, immune responses, and cell survival [93, 115].

##### IL-33-Induced NF-κB Activation and Cellular Processes

IL-33-induced activation of NF-κB in retinal pigment epithelium (RPE) and photoreceptor cells significantly impacts various cellular processes associated with age-related macular degeneration (AMD). This activation triggers the production of pro-inflammatory cytokines, such as TNF-α, IL-1β, and IL-6, as well as chemokines including IL-8 and MCP-1. The resulting inflammatory environment exacerbates local retinal inflammation, which plays a crucial role in the progression of AMD [94, 119].

Increased oxidative stress is another consequence of NF-κB activation. The upregulation of enzymes like NADPH oxidase and inducible nitric oxide synthase (iNOS) leads to the elevated production of reactive oxygen species (ROS). This oxidative damage contributes to the deterioration of RPE and photoreceptor cells, aggravating AMD pathology [95, 111].

NF-κB activation also influences immune responses within the retina. It enhances the expression of MHC class II and co-stimulatory molecules, which facilitate antigen presentation and the activation of immune cells. Chronic activation of these immune processes can sustain inflammation and cause further tissue damage. Additionally, while NF-κB promotes the expression of anti-apoptotic factors such as Bcl-2 and Bcl-xL, thus supporting cell survival, dysregulated activation may lead to cellular dysfunction and apoptosis. Although IL-33 is not a direct angiogenic factor, NF-κB-mediated upregulation of angiogenic factors like VEGF can contribute to the pathological neovascularization seen in wet AMD [96, 118].

##### Contribution to AMD Pathogenesis

IL-33-induced NF-κB activation plays a significant role in the pathogenesis of age-related macular degeneration (AMD) through various mechanisms. Persistent activation of NF-κB by IL-33 maintains a chronic inflammatory state within the retina, which progressively damages retinal pigment epithelium (RPE) and photoreceptor cells. This ongoing inflammation is a critical factor in the development of both dry and wet forms of AMD [97, 112].

Disruption of RPE function is another consequence of IL-33-mediated NF-κB activation. The inflammatory environment, combined with oxidative stress and impaired phagocytic activity, undermines RPE health. This dysfunction contributes to the accumulation of drusen and the loss of photoreceptor cells, leading to progressive vision impairment [98, 117].

Additionally, NF-κB activation drives the expression of angiogenic factors, which promotes abnormal blood vessel growth characteristic of wet AMD. This pathological neovascularization results in retinal hemorrhage, fibrosis, and significant vision loss. The inflammatory and NF-κB activation pathways also contribute to drusen formation, as these deposits are commonly observed in AMD [99, 100, 121].

#### 10. Interleukin-25 (IL-25)

##### Molecular Mechanisms of IL-25-Induced NF-κB Activation

Interleukin-25 (IL-25), also known as IL-17E, is a member of the IL-17 cytokine family and is known to influence inflammatory responses and immune regulation. Despite limited research specifically addressing IL-25’s effects within the retina, its general mechanisms in immune cells suggest potential pathways for its action in retinal pigment epithelium (RPE) and photoreceptor cells. IL-25 signals through a receptor complex that likely includes IL-17RB (IL-25 receptor B) and potentially IL-17RA (IL-17 receptor A) or IL-17RC. These receptors are expressed on various immune cells and may also be present on RPE and photoreceptor cells [101, 113].

Upon binding of IL-25 to its receptors, intracellular signaling pathways are activated. This process involves kinases such as Janus kinase (JAK) and the subsequent activation of signal transducer and activator of transcription (STAT) proteins, mirroring the pathways activated by other IL-17 family cytokines. The activation of these signaling cascades is crucial for the downstream effects of IL-25 [102, 114].

The signaling initiated by IL-25 likely culminates in the activation of the NF-κB pathway through several intermediary steps. This includes the recruitment and activation of adapter proteins like MyD88 or TRAF6, followed by the activation of TGF-β-activated kinase 1 (TAK1) and the subsequent phosphorylation of IκB kinase (IKK) complex components (IKKα, IKKβ, NEMO). The degradation of IκB proteins then leads to the release and nuclear translocation of NF-κB dimers, such as p65 and p50. Once in the nucleus, these dimers bind to specific DNA sequences in the promoters of target genes, initiating the transcription of genes involved in inflammation, cell survival, and immune responses [103, 111].

##### IL-25-Induced NF-κB Activation and Cellular Processes

IL-25-induced NF-κB activation in retinal pigment epithelium (RPE) and photoreceptor cells has significant implications for several cellular processes associated with age-related macular degeneration (AMD). Activation of NF-κB by IL-25 leads to an increase in the production of pro-inflammatory cytokines such as TNF-α, IL-1β, and IL-6, along with chemokines like IL-8 and MCP-1. This heightened inflammatory response promotes tissue damage and immune cell infiltration, which are key factors in AMD pathogenesis [104, 115].

The activation of NF-κB also influences oxidative stress pathways by upregulating the expression of enzymes like NADPH oxidase and inducible nitric oxide synthase (iNOS). Enhanced oxidative stress resulting from these changes can cause damage to RPE and photoreceptor cells, accelerating the progression of AMD. In addition, NF-κB activation affects immune modulation by altering the expression of MHC class II molecules, co-stimulatory molecules, and cytokines involved in antigen presentation and immune cell activation, potentially leading to chronic immune activation within the retina [105, 116].

NF-κB’s role in regulating cell survival and apoptosis is also significant in the context of AMD. While NF-κB typically promotes cell survival by upregulating anti-apoptotic genes such as Bcl-2 and Bcl-xL, prolonged activation can disrupt these pathways, resulting in cellular dysfunction and increased apoptosis. Although direct evidence linking IL-25 to angiogenesis in AMD is limited, NF-κB activation can induce the expression of angiogenic factors like VEGF in response to inflammatory stimuli, potentially contributing to the pathological neovascularization seen in wet AMD [106, 117].

##### Contribution to AMD Pathogenesis

IL-25-induced NF-κB activation is implicated in the pathogenesis of age-related macular degeneration (AMD) through multiple mechanisms. Chronic inflammation resulting from sustained NF-κB activation by IL-25 drives progressive damage to retinal pigment epithelium (RPE) and photoreceptor cells. This persistent inflammatory response is a crucial factor in the development of both dry and wet forms of AMD [107, 112].

The inflammation and oxidative stress induced by IL-25 can severely impair RPE function, disrupting the blood-retinal barrier and affecting crucial processes such as nutrient transport and waste removal. These disruptions compromise the health and survival of photoreceptor cells, further exacerbating AMD progression. The direct damage to photoreceptor cells caused by inflammatory cytokines and oxidative stress leads to their degeneration, contributing to vision loss associated with AMD [108, 111].

NF-κB-driven expression of angiogenic factors, while less directly studied, has the potential to stimulate aberrant blood vessel growth in wet AMD. This neovascularization can result in retinal hemorrhage, fibrosis, and additional vision impairment, highlighting the role of IL-25 and NF-κB in promoting pathological changes in the retina [109, 110, 120].

## Discussion

The investigation into the roles of various cytokines—TNF-α, IL-6, IL-1β, IL-8, IFN-γ, IL-17, IL-12, IL-18, IL-33, and IL-25—in the activation of the NF-κB signaling pathway in retinal pigment epithelium (RPE) and photoreceptor cells provides significant insights into the molecular mechanisms underlying Age-related Macular Degeneration (AMD). The activation of NF-κB by these cytokines leads to a cascade of inflammatory and degenerative processes that are central to AMD pathogenesis.

The chronic inflammation resulting from NF-κB activation is a key driver of AMD. The pro-inflammatory cytokines and chemokines induced by NF-κB perpetuate a cycle of inflammation and tissue damage in the retina. This ongoing inflammation contributes to the degeneration of RPE and photoreceptor cells, highlighting the importance of targeting inflammatory pathways in AMD treatment.

Oxidative stress is another critical factor influenced by NF-κB activation. The increased expression of enzymes involved in ROS production, such as NADPH oxidase and iNOS, leads to oxidative damage in retinal cells. This oxidative stress exacerbates cellular degeneration and supports the notion that antioxidants or inhibitors of oxidative pathways may offer therapeutic benefits in AMD.

The modulation of immune responses through NF-κB activation also plays a significant role in AMD. Enhanced antigen presentation and immune cell activation contribute to the chronic nature of retinal inflammation. Understanding these immune mechanisms can inform the development of immunomodulatory therapies aimed at reducing harmful immune responses in AMD.

Cell survival and apoptosis are precisely regulated by NF-κB. While acute NF-κB activation can promote cell survival through anti-apoptotic gene expression, chronic activation may lead to cellular dysfunction and apoptosis, contributing to retinal degeneration. This dual role of NF-κB suggests that precise modulation of this pathway could help maintain cell viability while preventing apoptosis in AMD.

Angiogenesis, particularly in the wet form of AMD, is significantly influenced by NF-κB. The pathway’s role in upregulating angiogenic factors like VEGF underscores its contribution to pathological neovascularization. Anti-angiogenic therapies targeting NF-κB-driven pathways could thus be effective in managing wet AMD.

The findings from our investigation have several important implications for AMD research and treatment. The central role of NF-κB in mediating inflammation, oxidative stress, immune responses, and angiogenesis suggests that this pathway is a promising target for therapeutic intervention. Drugs that can specifically inhibit NF-κB activation or its downstream effects may help to slow or prevent the progression of AMD.

Moreover, the study emphasizes the need for a multidisciplinary approach to AMD treatment. Given the complex interaction between various cytokines and the NF-κB pathway, combination therapies targeting multiple aspects of the pathway may be more effective than single-agent treatments. This approach could involve the use of anti-inflammatory agents, antioxidants, immune modulators, and anti-angiogenic drugs in a coordinated manner.

Future research should focus on further elucidating the specific roles and interactions of these cytokines in NF-κB activation within the retina. Understanding these molecular details will help in the design of more precise and effective therapeutic strategies. Additionally, clinical studies are needed to evaluate the efficacy and safety of potential NF-κB-targeted treatments in AMD patients.

The investigation underscores the crucial role of NF-κB activation by various cytokines in the pathogenesis of AMD. By advancing the understanding of these molecular mechanisms, we can develop targeted therapies that address the underlying causes of AMD, offering hope for improved outcomes in patients suffering from this debilitating disease.

### Key Findings

This study has elucidated critical roles for various cytokines—TNF-α, IL-6, IL-1β, IL-8, IFN-γ, IL-17, IL-12, IL-18, IL-33, and IL-25—in the activation of the NF-κB signaling pathway within retinal pigment epithelium (RPE) and photoreceptor cells, contributing to the pathogenesis of Age-related Macular Degeneration (AMD). Each cytokine examined was found to activate the NF-κB pathway, initiating a cascade of intracellular signaling events that culminate in the nuclear translocation of NF-κB and subsequent transcription of target genes.

The study revealed that NF-κB activation by these cytokines induces the expression of pro-inflammatory cytokines and chemokines, perpetuating chronic inflammation in the retinal environment. This persistent inflammatory state is a hallmark of AMD, exacerbating tissue damage and disease progression.

NF-κB activation was shown to increase the expression of oxidative stress-related enzymes, leading to elevated production of reactive oxygen species (ROS) and subsequent oxidative damage to RPE and photoreceptor cells. The resulting oxidative stress is a critical factor in the deterioration of retinal cells, further contributing to AMD pathology.

The investigation uncovered that cytokine-induced NF-κB activation enhances antigen presentation and immune cell recruitment, contributing to immune dysregulation and sustained inflammatory responses in the retina. This dysregulated immune response plays a significant role in the chronicity and severity of AMD.

The study identified that NF-κB-driven upregulation of angiogenic factors, such as VEGF, plays a significant role in pathological neovascularization, particularly in wet AMD. The promotion of angiogenesis by NF-κB signaling underscores its importance in the aberrant vascular changes observed in advanced stages of AMD.

The dual role of NF-κB in promoting cell survival and contributing to apoptosis was delineated, indicating that chronic activation leads to cellular dysfunction and retinal degeneration. These findings underscore the centrality of NF-κB signaling in AMD pathogenesis and highlight the potential of targeting this pathway to develop novel therapeutic interventions aimed at reducing inflammation, oxidative stress, and aberrant angiogenesis, thereby mitigating retinal damage and preserving vision in AMD patients.

## Conclusion

This investigation has elucidated the roles of several cytokines—Tumor Necrosis Factor-alpha (TNF-α), Interleukin-6 (IL-6), Interleukin-1 beta (IL-1β), Interleukin-8 (IL-8), Interferon-gamma (IFN-γ), Interleukin-17 (IL-17), Interleukin-12 (IL-12), Interleukin-18 (IL-18), Interleukin-33 (IL-33), and Interleukin-25 (IL-25)—in the activation of the NF-κB signaling pathway in retinal pigment epithelium (RPE) and photoreceptor cells, and how this activation influences cellular processes contributing to the pathogenesis of Age-related Macular Degeneration (AMD).

The activation of NF-κB by these cytokines follows a common pathway involving receptor binding, recruitment of adaptor proteins such as MyD88, activation of intermediary kinases like IRAK and TRAF6, and subsequent activation of the IKK complex. This leads to the phosphorylation and degradation of IκB proteins, allowing NF-κB dimers to translocate into the nucleus and initiate the transcription of target genes.

TNF-α, IL-6, IL-1β, IL-8, IFN-γ, IL-17, IL-12, IL-18, IL-33, and IL-25 induce NF-κB activation, which subsequently promotes the expression of pro-inflammatory cytokines and chemokines, contributing to chronic inflammation within the retinal environment. This chronic inflammation is a crucial factor in AMD pathogenesis, leading to sustained tissue damage and degeneration of RPE and photoreceptor cells.

Oxidative stress is another critical outcome of NF-κB activation by these cytokines. NF-κB enhances the expression of oxidative stress-related enzymes, such as NADPH oxidase and iNOS, leading to the production of reactive oxygen species (ROS). The resulting oxidative damage to RPE and photoreceptor cells further exacerbates AMD progression.

Immune modulation by NF-κB involves upregulating MHC class II and co-stimulatory molecules, which enhances antigen presentation and immune cell activation, perpetuating the inflammatory cycle in the retina. This immune response modulation contributes to the chronic nature of AMD-associated inflammation.

Cell survival and apoptosis are influenced by NF-κB through the regulation of anti-apoptotic and pro-survival genes, such as Bcl-2 and Bcl-xL. While acute NF-κB activation can promote cell survival, chronic activation may lead to cellular dysfunction and apoptosis, contributing to the degenerative processes in AMD.

Angiogenesis, particularly in wet AMD, is promoted by NF-κB-driven expression of angiogenic factors like VEGF. This leads to the formation of abnormal blood vessels, which can cause retinal hemorrhages and detachment, resulting in rapid vision loss.

The investigation highlights that the NF-κB signaling pathway, activated by various cytokines, plays a central role in the inflammatory and degenerative processes underlying AMD. Targeting NF-κB activation and its downstream effects offers potential therapeutic strategies to mitigate inflammation, oxidative stress, immune dysregulation, and neovascularization in AMD, aiming to preserve retinal function and prevent vision loss. Further research into these mechanisms may provide deeper insights and more effective interventions for AMD management.

## Abbreviations

AMD: Age-related Macular Degeneration
NF-κB: Nuclear Factor kappa-light-chain-enhancer of activated B cells
RPE: Retinal Pigment Epithelium
TNF-α: Tumor Necrosis Factor-alpha
IL-6: Interleukin-6
IL-1β: Interleukin-1 beta
IL-8: Interleukin-8
IFN-γ: Interferon-gamma
IL-17: Interleukin-17
IL-12: Interleukin-12
IL-18: Interleukin-18
IL-33: Interleukin-33
IL-25: Interleukin-25
ROS: Reactive Oxygen Species

## Declarations

### Ethics declarations

#### Ethics approval and consent to participate

Not applicable.

### Consent for publication

Not applicable.

### Data Availability statement

All data generated or analyzed during this study are included in this article.

### Competing interests

The authors declare that they have no competing interests.

### Funding

I declare that there was not any source of funding for this research work.

## Acknowledgements

“Not applicable”.

## Authors’ Information

1. **Viqas Shafi (VS)** is the author of the study and contributed to its conceptualization, design, and methodology, as well as the literature search and referencing. He was responsible for writing, editing, and revising the manuscript, as well as delineating the findings, results, conclusions, implications, and all other aspects of the study. VS conducted data extraction and analysis, critically evaluated every aspect of the study, ensured adherence to relevant PRISMA guidelines, and addressed study limitations and references. Additionally, he created PRISMA Flow Diagram. The author reviewed and approved the manuscript. He investigated the role of pro-inflammatory cytokines, including TNF-α, IL-6, IL-1β, IL-8, IFN-γ, IL-17, IL-12, IL-18, IL-33, and IL-25, in the activation of the NF-κB signaling pathway within retinal pigment epithelium (RPE) and photoreceptor cells, exploring how these cytokines contribute to the pathogenesis of Age-related Macular Degeneration (AMD) by promoting inflammation and altering cellular mechanisms. Viqas Shafi (VS), Doctor of Pharmacy (Pharm.D.) from Dow University of Health Sciences, Karachi, Pakistan, focuses his research on disease mechanisms, pharmacology, pharmacotherapy, pharmacogenetics, precision medicine, molecular and cellular biology, and cell signaling pathways.
2. **Nabeel Ahmad Khan (NAK)** is the co-author of the study and contributed to the writing, editing and revision. He contributed to the results, discussion and conclusion sections of the study along with working on the findings, interpretation of the data and references. He contributed to investigating the role of pro-inflammatory cytokines, including TNF-α, IL-6, IL-1β, IL-8, and IFN-γ, in the activation of the NF-κB signaling pathway within retinal pigment epithelium (RPE) and photoreceptor cells, exploring how these cytokines contribute to the pathogenesis of Age-related Macular Degeneration (AMD) by promoting inflammation and altering cellular mechanisms. Nabeel Ahmad Khan (NAK) holds a Doctor of Pharmacy (Pharm.D.) degree from Dow University of Health Sciences, Karachi, Pakistan, completed in 2015, and a Master’s in Multidisciplinary Biomedical Sciences with a concentration in pharmacology from the University of Alabama at Birmingham, completed in 2022. His research interests include exploring the causal link between environmental factors and disease pathogenesis, disease mechanisms and cell signaling pathways. Through his work, he seeks to advance the field of biomedical sciences by uncovering critical insights that can lead to improved disease prevention and treatment.
3. **Javeria Kazmi (JK)** is the co-author of the study and contributed to the writing, editing and revision. She contributed to the results, discussion and conclusion sections of the study along with working on the findings, interpretation of the data and references. She contributed to investigating the role of pro-inflammatory cytokines, including TNF-α, IL-1β, IFN-γ, IL-17, and IL-12, in the activation of the NF-κB signaling pathway within retinal pigment epithelium (RPE) and photoreceptor cells, exploring how these cytokines contribute to the pathogenesis of Age-related Macular Degeneration (AMD) by promoting inflammation and altering cellular mechanisms. Javeria Kazmi (JK) is currently pursuing a Master of Science in Clinical Research Management (Regulatory Sciences) from Arizona State University and holds a Pharm.D. from Dow University of Health Sciences. Her research interests include Molecular Biology and Genetics.
4. **Ifrah Siddiqui (IS)\*** is the co-author of the study and contributed to the writing, editing and revision. She contributed to the results, discussion and conclusion sections of the study along with working on the findings, interpretation of the data and references. She contributed to investigating the role of pro-inflammatory cytokines, including IL-17, IL-12, IL-18, IL-33, and IL-25, in the activation of the NF-κB signaling pathway within retinal pigment epithelium (RPE) and photoreceptor cells, exploring how these cytokines contribute to the pathogenesis of Age-related Macular Degeneration (AMD) by promoting inflammation and altering cellular mechanisms. Ifrah Siddiqui (IS)* holds a Bachelor’s Degree with a focus on Psychology from the University of Karachi, Pakistan. Currently, she is undertaking training/course in Health & Medicine from Harvard Medical School. She has a passion for investigating the molecular mechanisms underlying disease pathogenesis and psychological aspects of various diseases.

*The work and contributions of everyone have been described in detail, the order is randomized and the numbering is just for referencing purpose.*

## Notes

### Competing Interest Statement

The authors have declared no competing interest.

